# Aggresssion in School Children: Role of Gender, Family Factors and Exposure to Violence

**DOI:** 10.1101/2021.03.25.21254374

**Authors:** Mukul Sehgal, Ajita Nayak

## Abstract

**INTRODUCTION:** Increasing instances of aggression in school children has become a cause of great concern for educationists, parents and mental health professionals. With increased globalization and information overload, today’s child is exposed to influences which probably adversely modify the child’s behavior. These aggressive tendencies could affect the child’s academic, social functioning and also may lead to psychiatric problems. Hence, this study was planned to find out the amount of aggression among school children and various potential contributing factors for the aggressive behavior. Identifying these risk factors could help design preventive strategies among school children.

**OBJECTIVE:** To evaluate the amount of aggression and its contributing factors among 10-12 year-old children.

**SUBJECTS:** The subjects involved the school children and their teachers in various schools across Mumbai. The age group of study was 10-12 years i.e. class 5^th^ to class 8^th^

No. of subjects:-102

**STUDY PROCEDURE:** A total of 102, 10-12 year-old school children (5th to 8th class) in the Mumbai metropolitan area were interviewed to assess the presence of aggression. A validated scale, Children’s Aggression Scale – Teacher version (CAS-T), was used to assess the presence of aggression. Children were assessed & scored on basis of this scale with questions answered by their teachers. Data was analyzed on the basis of each factor using unpaired t-test, ANOVA test and Kruskal-Wallis Test (Nonparametric ANOVA). Institutional ethics committee approval was taken for research at Seth GS Medical College, Mumbai, India.

**RESULTS:** A high level of aggression was present in 4% other children while moderate levels were present in 8%. Boys were found to be more aggressive than girls (p-value 0.003). Aggression scores were significantly higher in children from English medium and with poor academic performance. Children exposed to physical abuse and violence on Television showed significantly high aggression. Perceived parental conflict in boys was found to be significantly associated with lower aggression scores. Among the various factors studied, aggression scores showed highest correlation with exposure to violence on television for more than 2 hours/day (11.67 vs 3.19 p-value <0.001).

**CONCLUSION:** About 12% of schoolchildren in the 10-12years group show aggressive behavior needing interventions. Boys from abusive families, with poor academic performance and exposed to excessive violence on TV seem particularly at risk for developing aggressive behavior.

## INTRODUCTION

We have lately seen an increasing amount of incidence of aggression among adolescents and young adults.^1^ It becomes even more important to evaluate the causes of aggression. Experiences during early childhood and adolescence have powerful and enduring effect on physiological and psychological development and, therefore on their behavior. There is a vast literature suggesting that early maltreatment of children is associated with the development of aggressive and maladaptive behavior.^2-4^ Whereas complete emotional deprivation of infants leads to severe depression and restriction of growth and development, the lesser degree of neglect has been associated with poor peer relationships and aggressive behaviors.^5^ Severe physical abuse has been associated with subsequent extremely violent behavior.^6^ Maltreatment affects every aspect of functioning-cognitive, emotional and physiological.^7^ Lately there has been an increase in use of internet among adolescents. According to a study published in *New Media and Society Journal (2015)*, the probability of using smartphones by younger children is increasing, and more so for more affluent families.

Children, in modern day society, are very vulnerable to the media which causes a deep rooted effect on the behavior of a child.^8^ The very quality of aggression depicted in the media has changed over the past several years, becoming increasingly graphic.^9^ There is evidence that suggests a violent environment affects the child’s behavior negatively.^10,11^ Small children seem to react differently from older children to observing the effects of aggression among others.^12^ Younger children tend to choose violent outcomes for violent films, whereas older children while choosing an outcome take into consideration whether or not the violence they had observed at the beginning of the film are justified or not. Behavioral problems are sometimes associated with multi-organ dysfuctions later in life including cardiovascular disease, sepsis, various syndromes and other infections.^13-18^

It is hypothesized that violence and aggression would increase in societies when social support systems malfunctioned or totally failed. Physical pain, discomfort, Neurological dysfunction, abusive upbringing, rates of mental illness are more pervasive in these disadvantaged communities and contributes to disproportionately higher rate of violence found in these social situations.

The programs, which enhance emotional stability, encourage a self-control, improve judgment, promote self-esteem, and reinforce a sense of security, develop the ability to recognize one’s own feelings and the feelings of others, and increase the ability to express these feelings verbally will raise the threshold for aggression and diminish the likelihood of violent behavior.^19^ The obvious settings in which to begin to attempt to promote these enhancers of civilized interactions are homes and schools.

However, the data on the aggressive behaviors among school children in India and their causes is lacking. We could use the knowledge of the causes of violence to create systems that would have the opposite effects. Our knowledge about psycho-physiological correlates of civilized behaviors could have important implications for reducing violence by modifying the ways in which we organize our everyday lives. Hence this study was designed to identify the aggression in school children and factors associated with it. The objectives of this were to assess level of aggression in 10-12 year-old school children in the Mumbai Metropolitan area and the factors that affect aggressive behavior.

## AIMS & OBJECTIVES

1. To study and assess aggression in school children.
2. To study the association of aggression with socio-demographic profile, medium of education, academic performance, school attendance, leisure activities, exposure to violence on television, perceived parental conflict and history of physical abuse.

## METHODOLOGY

The study was performed after obtaining the permission of Institutional Ethics Committee of Seth GS Medical College and KEM Hospital, Mumbai. A written informed consent was taken from the parent/ guardian of the children before enrollment in the study. Fifth to 8th class school children, aged 10-12 years and their teachers were enrolled in the study. Frequent absentees and those who did not provide consent were excluded.

### STUDY PROCEDURE

Students were selected randomly from different classes mostly from Grade 5^th^ to 8^th^ from 2 different schools in Mumbai, India (one of them had English as medium of teaching another was in vernacular medium). The students were selected based on random roll numbers (based on random no. figure). Informed consent was obtained from parents of selected students (based on inclusion and exclusion criteria). Students were then interviewed on the basis of Pre-determined proforma that contained the questions regarding Psycho-Social correlates being studied. As television formed the primary source of media exposure, it was used to determine the effect of violence in media. The school performance and attendance was obtained from their class teachers. The class teachers were asked to answer the CAS-T scale (Children’s Aggression Scale – Teacher version) about the same students, to assess the level of aggression these children.

### MATERIALS USED

1) Semi-structured Proforma to note socio demographic details, academic performance, attendance, time spent in leisure activity, time of exposure to violence on television, perceived parental conflict and history of physical abuse.

2) CAS-T (Children’s Aggression Scale – Teacher version)

CAS-T comprises of 23 items. These items assess 4 domains which include physical aggression, verbal aggression, aggression against objects and animals and use of weapons. The items are rated on 5-point scale with responses: never, once/month or less, once/week or less, 2-3 times/week, and most days or with responses: never, once a week, 3-5 times, 5-10 times, and more than 10 times.

3) AlsoGraphPadInStat 3.10 was used to analyze Kruskal-Wallis Test (Nonparametric ANOVA).

### STATISTICAL ANALYSIS

t-test was used for the analysis of two variables. ANOVA test was used to compare more than two independent variables. Post-hoc test was used to analyze results of ANOVA test of Normality was done before ANOVA to find the distribution of data.

Kruskal-Wallis Test (Nonparametric ANOVA) to assess data when normality of one or more variables is not as per Gaussian curve. Since all the data did not pass the test of normality, the results of ANOVA test although statistically significant were not relied upon. Therefore it was decided to use Kruskal-Wallis Test (Nonparametric ANOVA).

## RESULTS

A total of 102 students were included in the study. The sample consisted of 102 students. Of these, 43 were boys and 59 were girls. 47% of the students studied in the vernacular medium while 53% in the English medium. The social condition include, 66% of students are from the nuclear family while 34 % students belonged to the joint or extended family. All the children belonged to the middle socioeconomic class.

The school performance and attendance were obtained from the teachers. It was found that 91% of students had regular attendance while 9% of students were irregular. Good school performance (defined as >75% aggregate score in Annual exam) was present in 34% students.

Using the Children’s Aggression Scale-Teacher’s version (CAS-T), the teachers were asked to rate the students on physical aggression, verbal aggression, aggression against objects and animals and use of weapons. Individual scores on these parameters and total scores on CAS-T were calculated. The total scores were divided into low, moderate and high based on the maximum score obtained. High scores were present in 4% of the sample and 8% of the children were found to have moderate scores. In 88 % of the children, low levels of aggression was obtained (Figure 1 & 1A).

**Figure 1.**
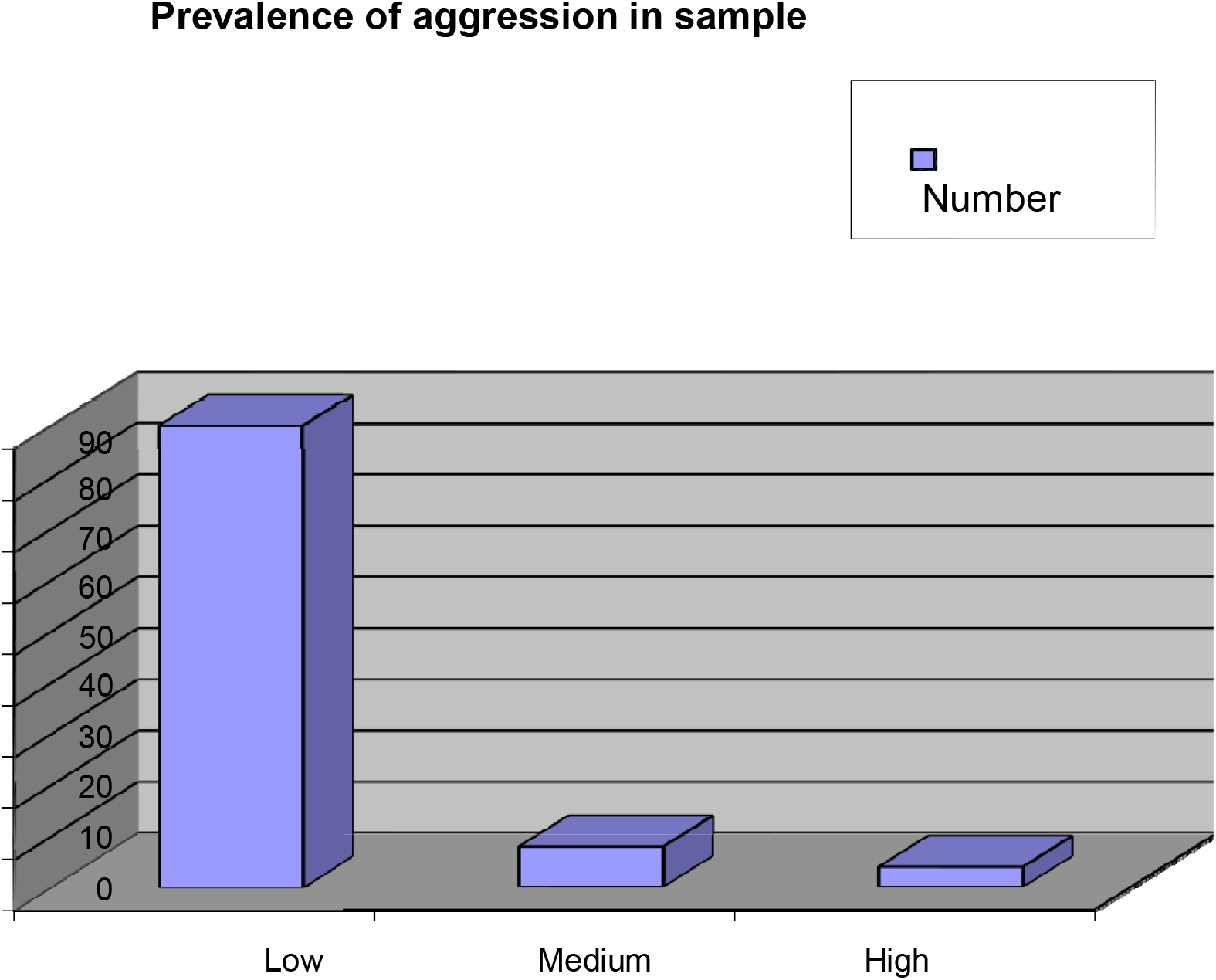
Prevalence of aggression in sample. Low level of aggression based on CAS-T score is found in 90 children as compared to medium and high aggression in 8 and 4 kids respectively. Both Physical and verbal aggression almost show the same pattern in distribution among the sample of our study, but still the children are more likely to have high prevalence of physical aggression (2.95%) as compared to verbal aggression (0.98%)

T-test was used to compare various socio-demographic, academic and family environment variables. The aggression scores were significantly higher in boys when compared to the girls (p<0.05) (Figure 2). Aggression was also higher in children coming from nuclear families (m4.28 ± SD) as compared to those from joint families (3.94±SD) (Figure 3).The children were also grouped into two based on their academic performance. Lower aggression scores showed significant correlation with good academic performance (p <0.05) (Figure 4). However no significant correlation was found with the attendance (Figure 5). Children from the vernacular medium of education had significantly lower scores on aggression (Mean-2.76) as compared to children from the English medium (Mean-5.36) (Figure 6).

**Figure 2.**
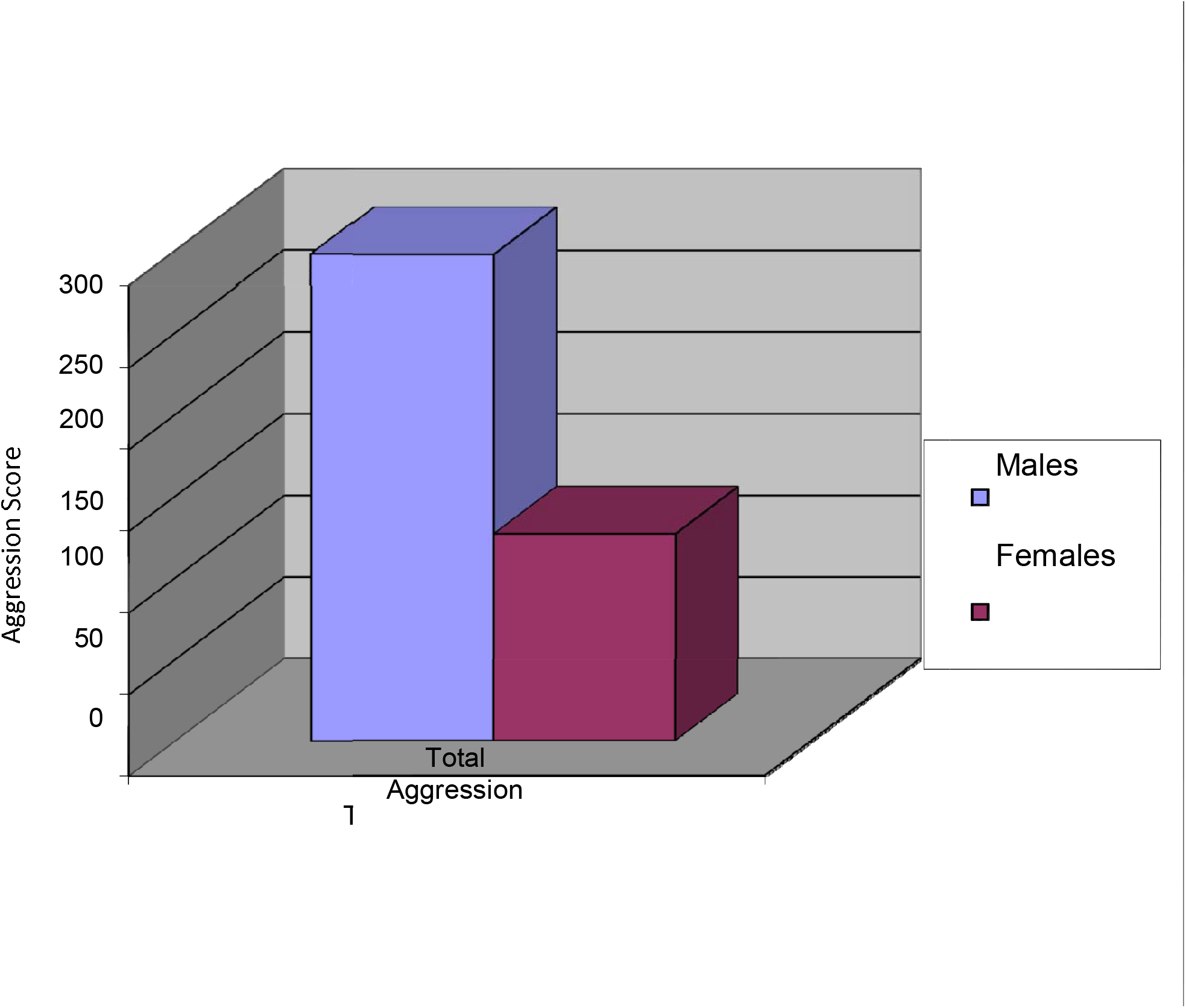
Gender differences in aggression score When compared on basis of Gender the aggression was found to be significantly (p=0.0003) higher in males (mean=6.93) as compared to females (mean=2.15).

**Figure 3.**
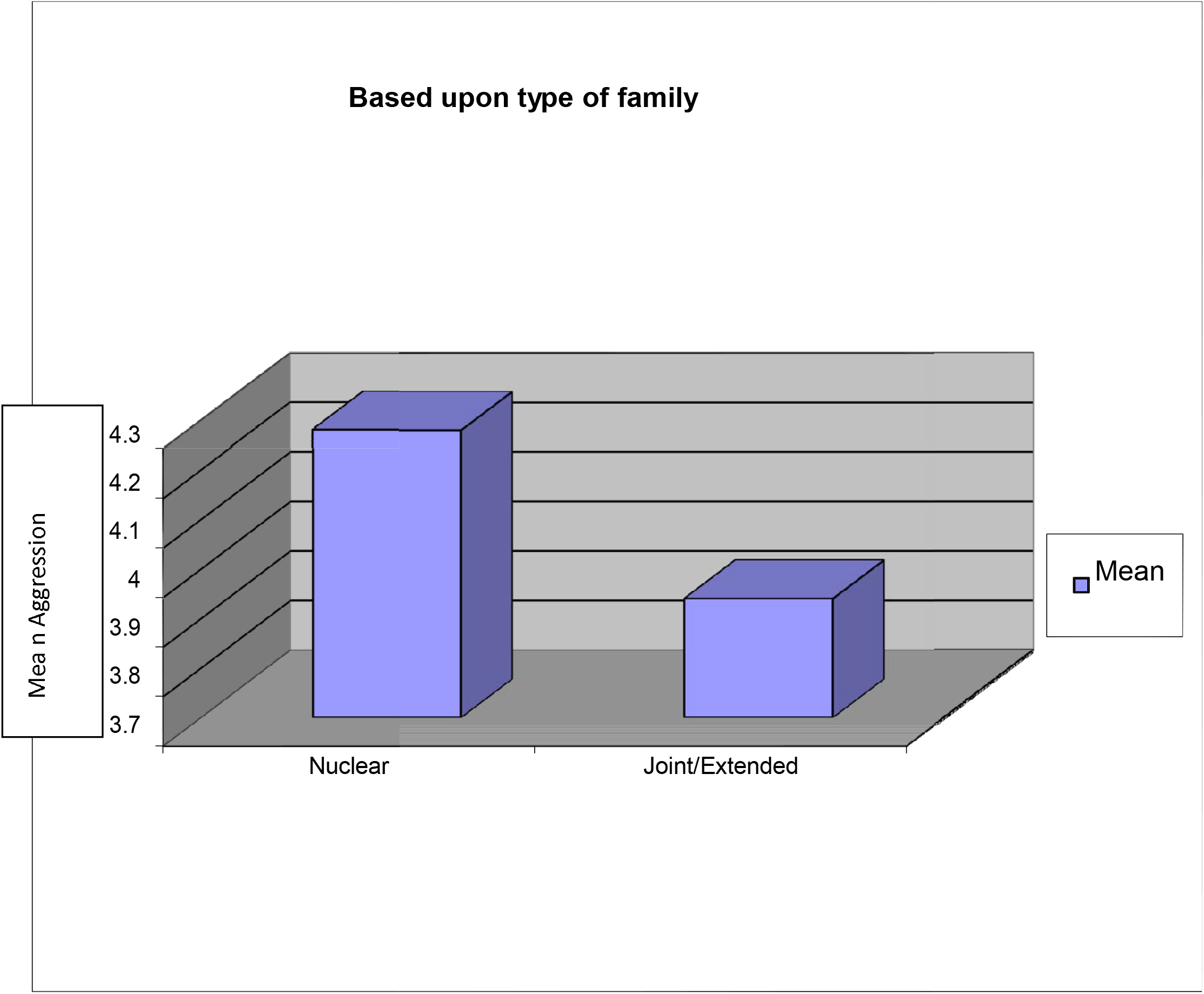
Comparison of aggression based upon type of family Although aggression scores are higher inn nuclear as compared to joint family, but the difference is not statistically significant (p=0.8).

**Figure 4.**
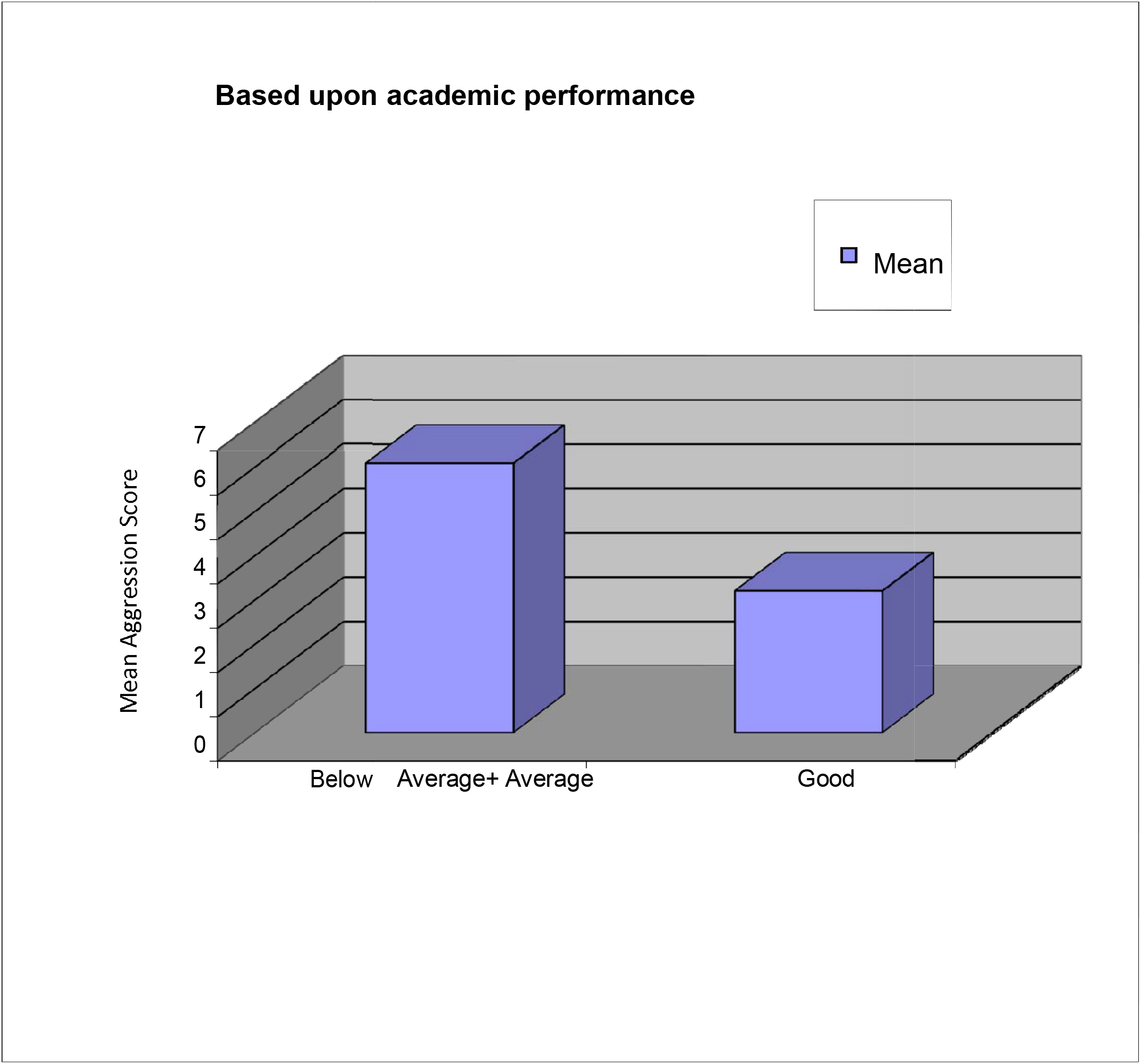
Comparison of aggression based upon academic performance. It was found that children who did well in school have lower level of aggression (Mean=6.08) than their peers (mean= 3.2) which was indeed statistically significant. (p=0.041)

**Figure 5.**
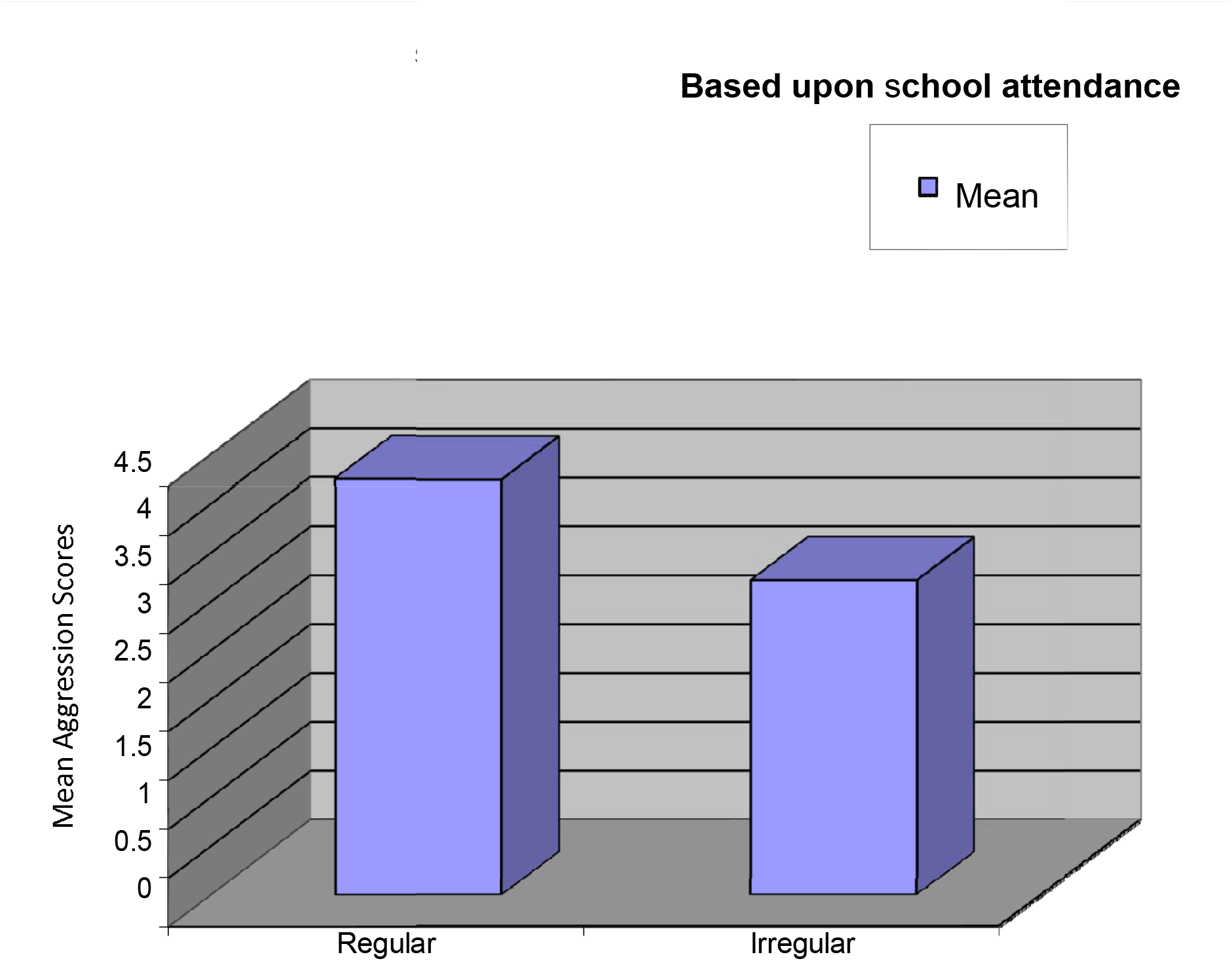
Comparison of Aggression Scores based on school attendance. Although scores are higher in those with regular attendance it is not Statistically (p=0.66).

**Figure 6.**
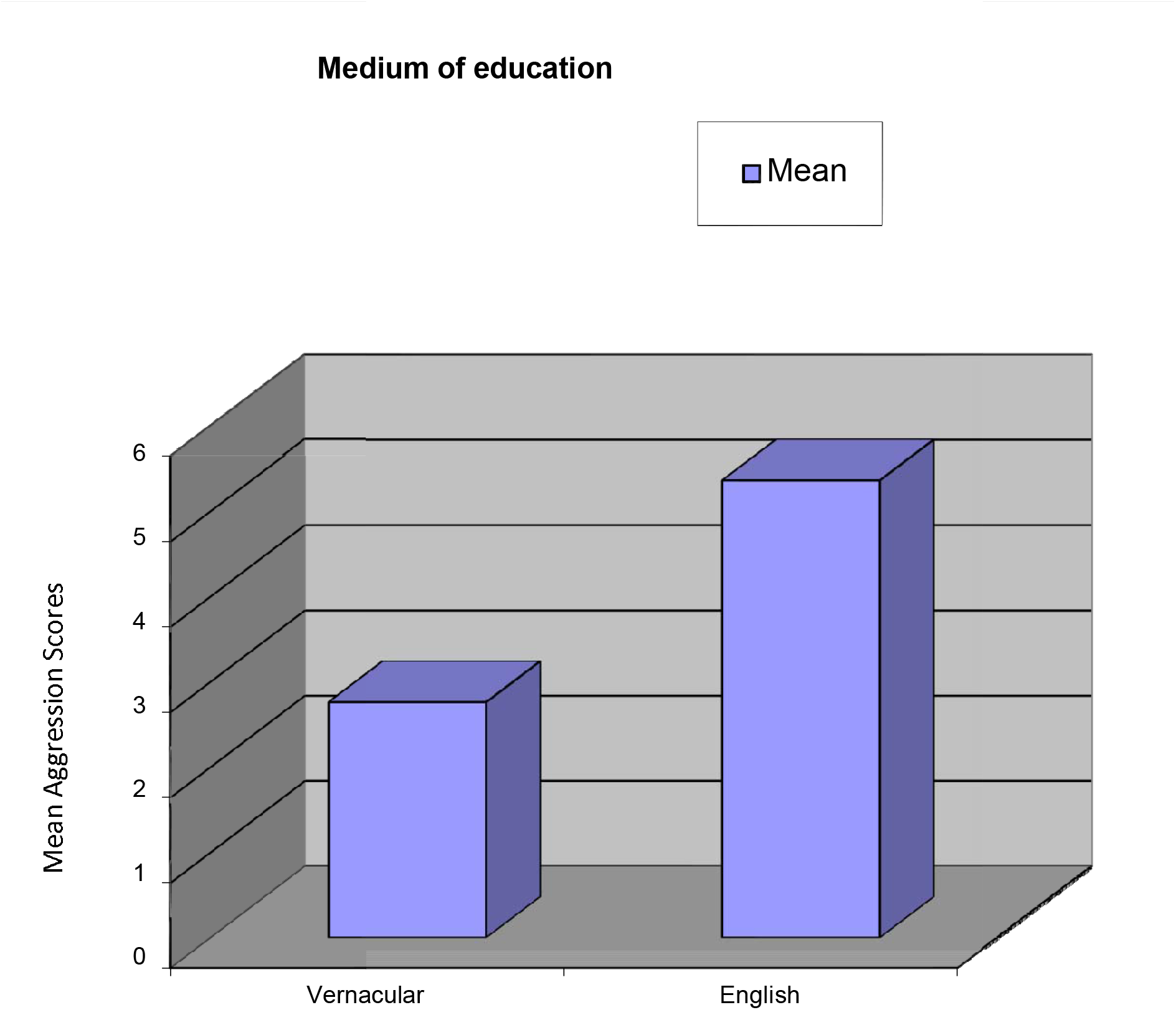
Comparison of Aggression Scores based upon medium of education. Students from English medium schools were found to have higher aggression scores (mean= 5.36) as compared that of vernacular medium (mean=2.76), which was statistically significant (p=0.05).

The association between history of physical abuse and perceived parental conflict and aggression scores was also studied. Significantly higher scores were obtained in children with physical abuse (p<0.05) (Figure 7). Perceived parental conflict was however not significantly associated with higher aggression scores (Figure 8).

**Figure 7.**
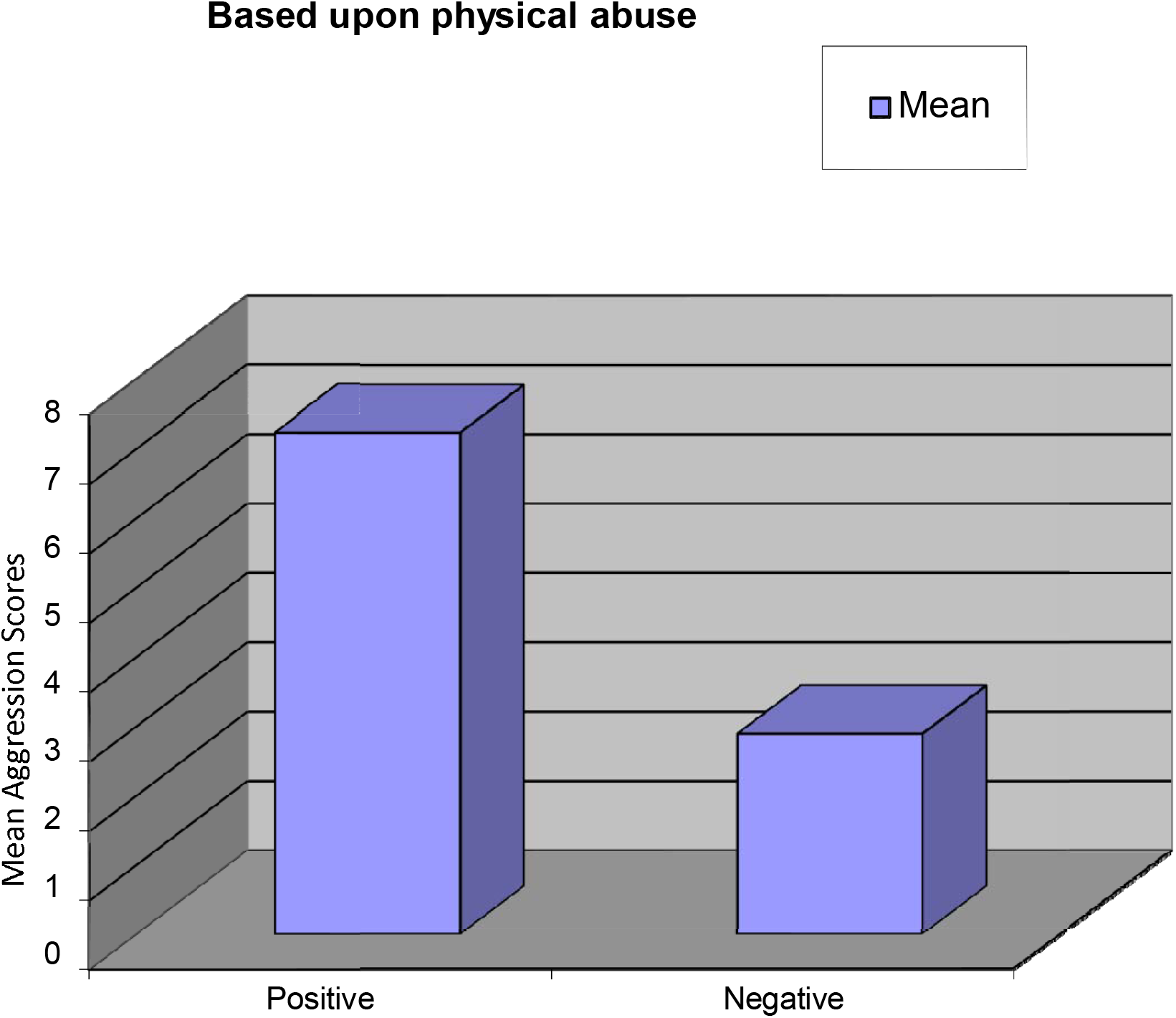
Comparison of aggression scores based upon history of physical abuse. It is found that those that have positive history have higher total aggression scores (mean=7.23) than those without it (mean=2.89), which is statistically significant (p=0.002).

**Figure 8.**
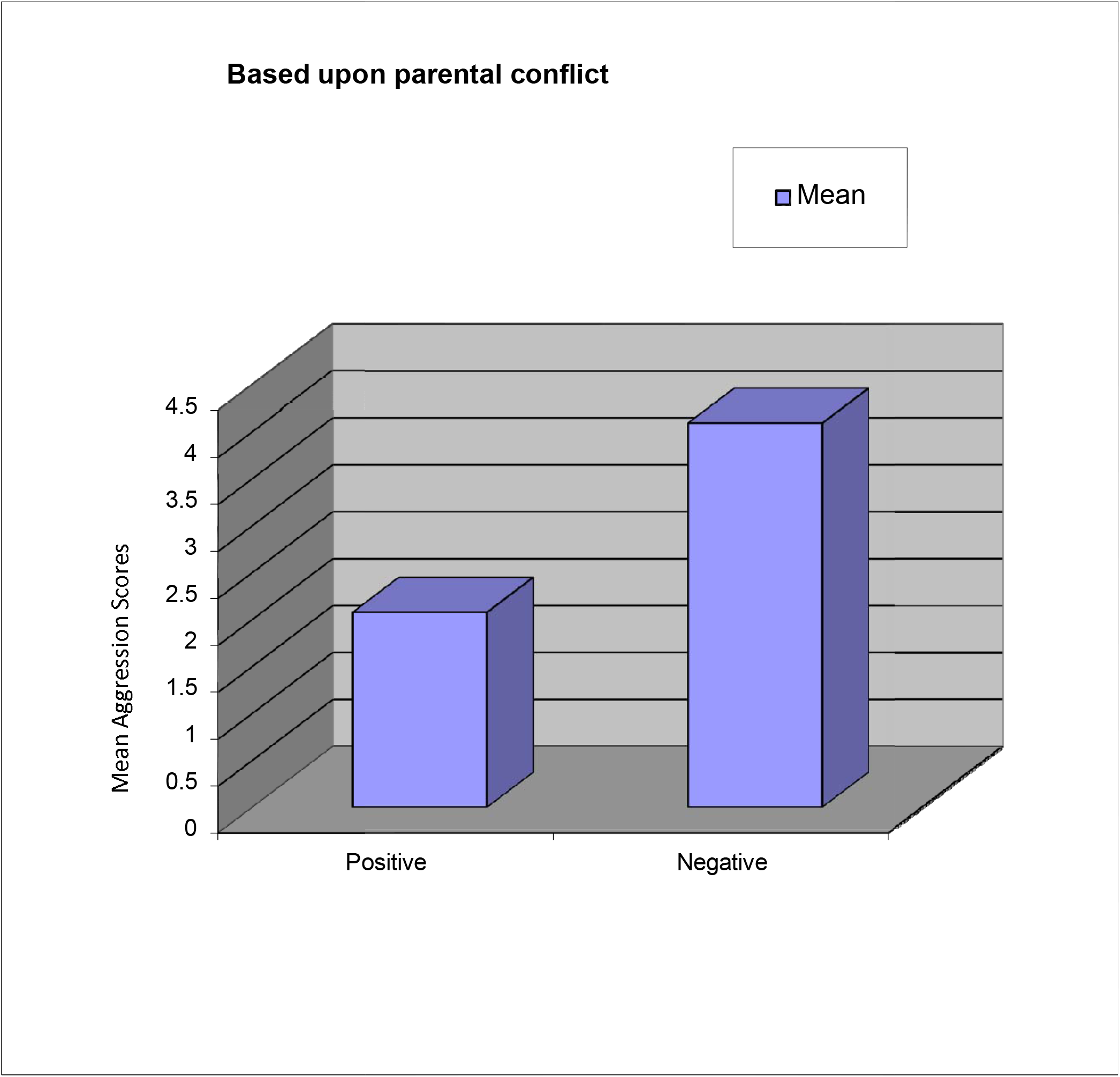
Comparison of aggression scores based upon history of perceived parental conflict. Although those children who perceived parental conflict have higher aggression scores but these results are not statistically significant (p=0.76)

An average of 2 hours/day of play time and television watching is seen in Indian children. It was found that aggression scores were significantly higher in children exposed to >2 hours of violent TV (Figure 9) .Those who spent >2 hours in play also had significantly higher scores (Figure 10).

**Figure 9.**
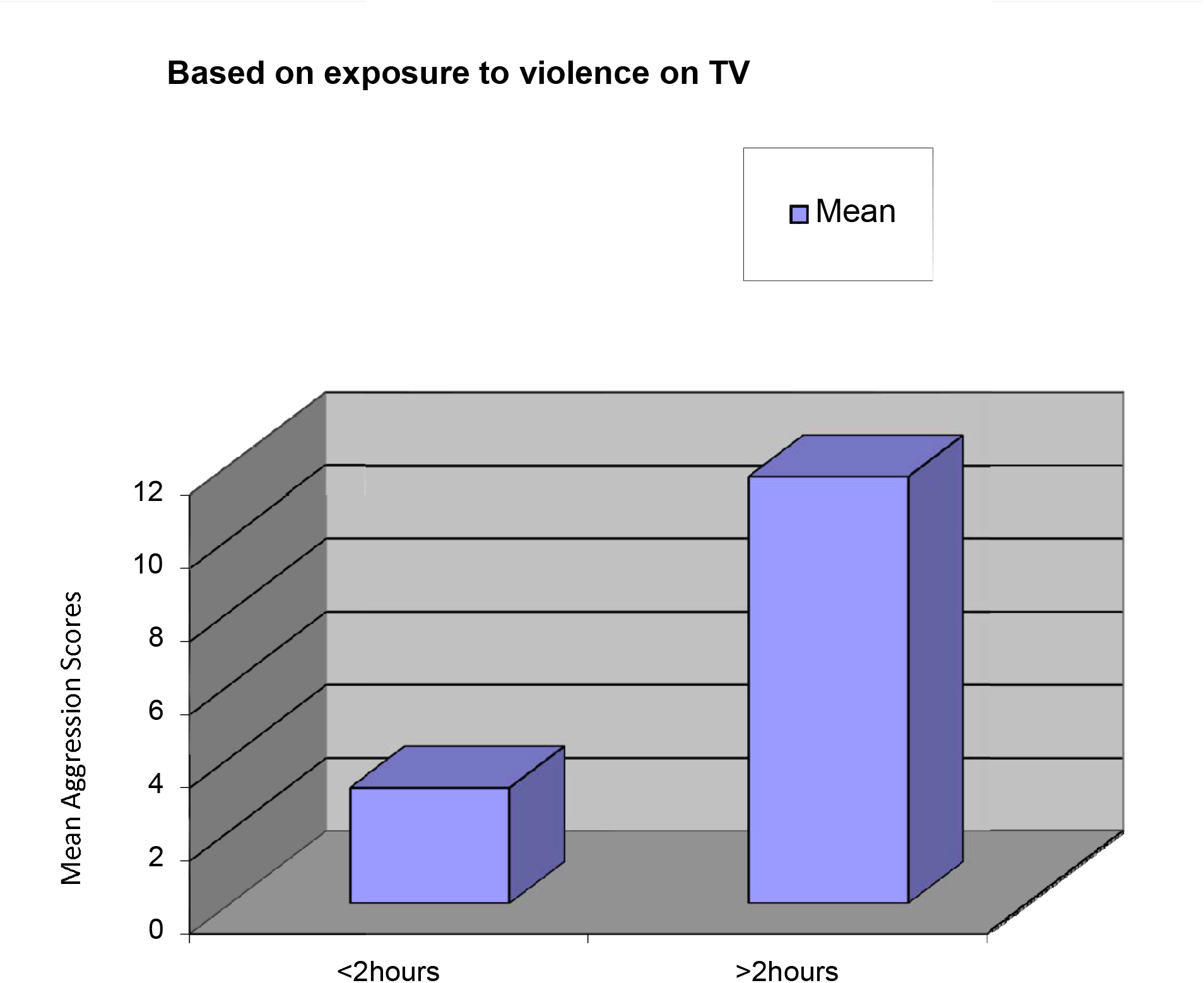
Comparison based upon exposure to violence on T.V. It was found that aggression scores are higher in those who were exposed to violence in TV for more than 2hrs/day (mean=11.67) than those watched less than 2hrs/day (mean=3.16) which was statistically significant. (p=0.000003)

**Figure 10.**
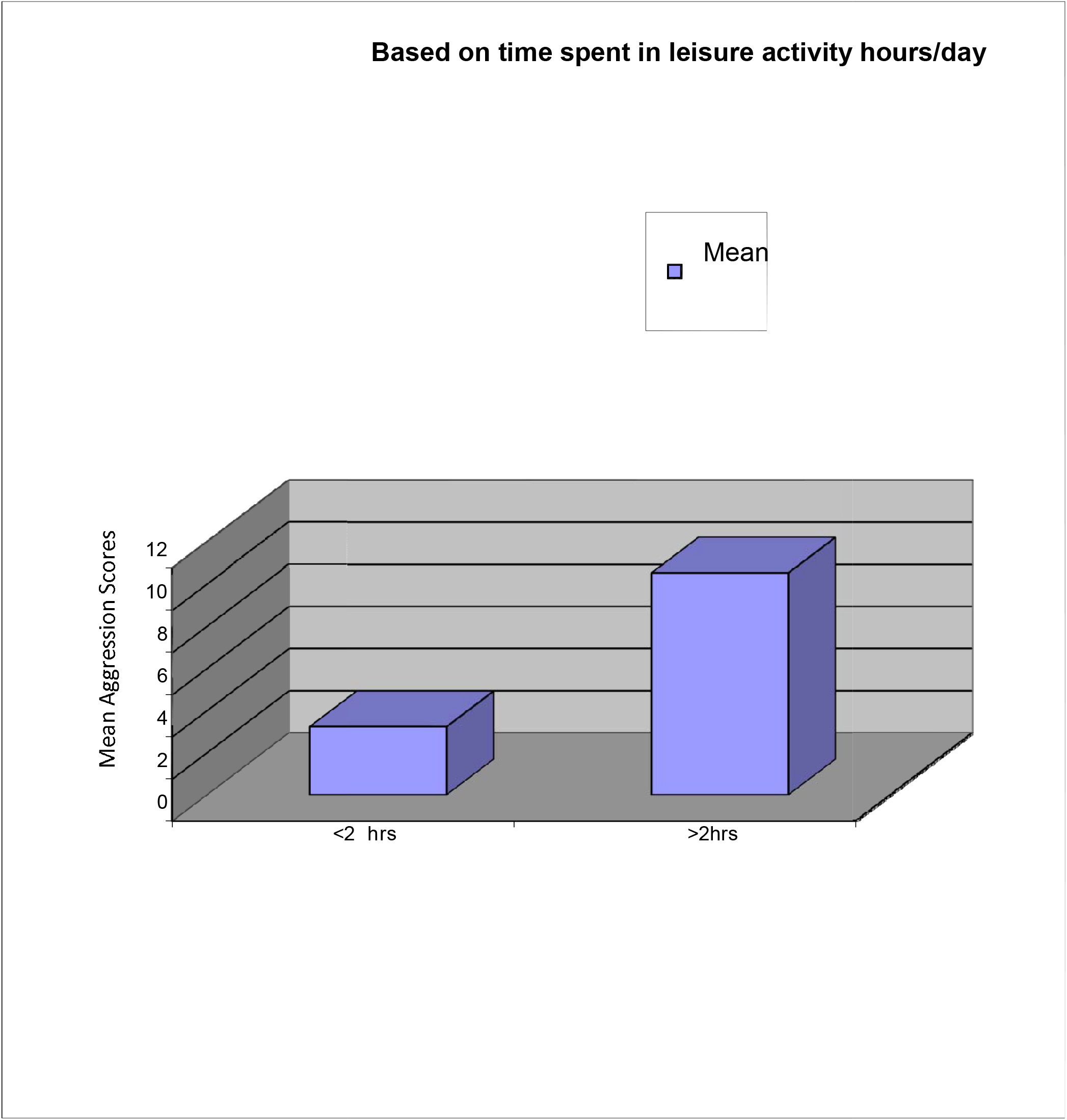
Comparison of aggression scores based upon time spent in leisure activities. Those who spent time in leisure activities for more than 2 hrs/day had higher aggression (mean=10.53) than those who spent less than 2hrs/day (mean=3.23), which is statistically significant. (p=0.00018)

When groups were divided on basis of gender and amount of exposure to violence on TV (6 groups), it was found that (Figure 11) boys who watched violent content on TV for more than 2hours/day had highest aggression which was found statistically significant by ANOVA test. (F >F_critical_).

**Figure 11.**
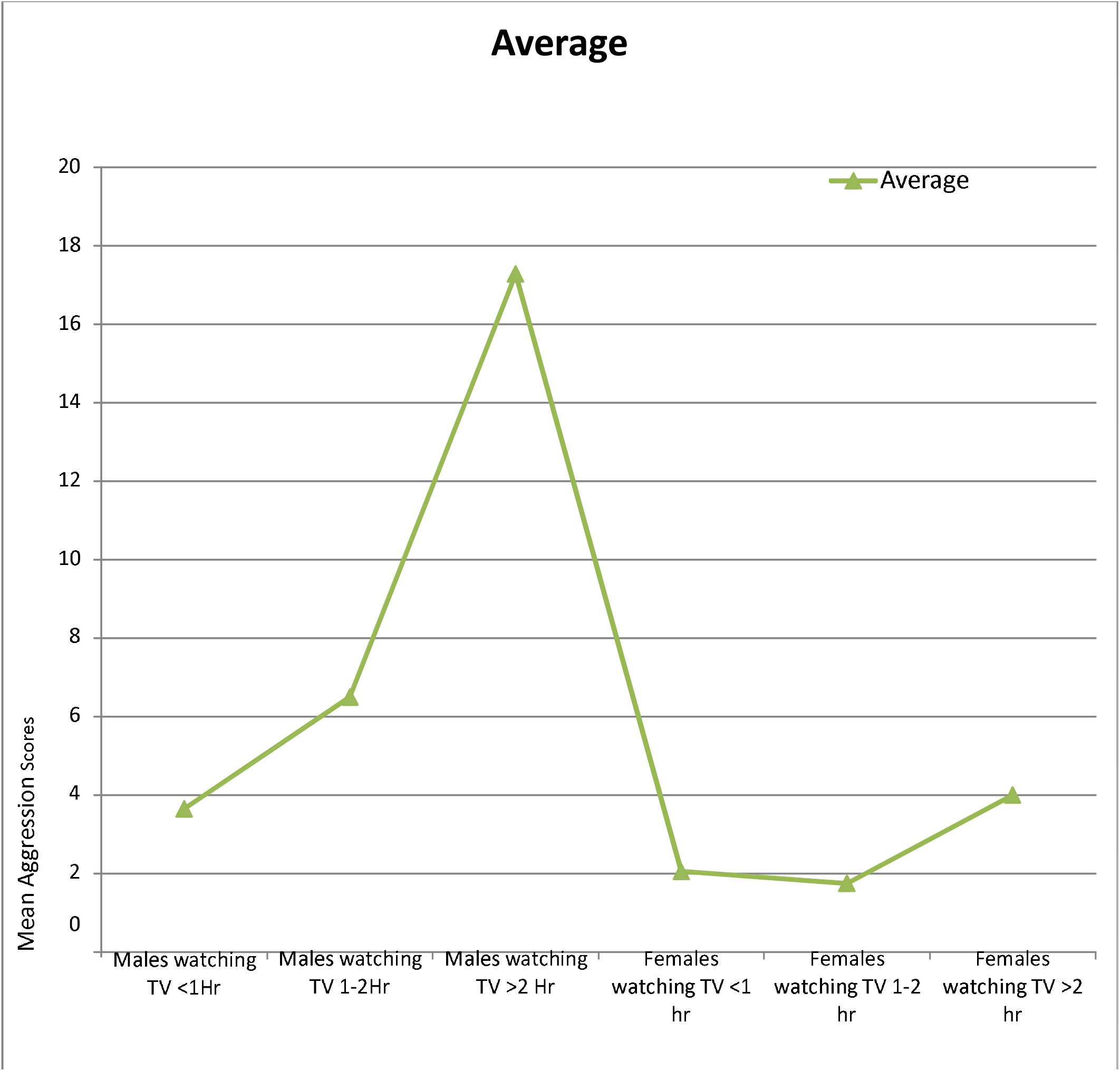
Comparison based upon Exposure to violence on T.V using ANOVA tests and comparing various sub-groups of population. It was found that aggression scores were highest among males those were exposed to violence on TV for more than 2hrs/day. (F=9.06, F_critical_=2.31 & p=4.53 × 10^−7^). Since, F >F_critical_ at p=0.05 therefore, this difference is more than that expected by chance.

Similarly, although there was no significant difference found in children with and without parental conflict, the difference was significant when data was sorted out on gender basis and analyzed by ANOVA test. It was found that boys without parental conflict had more aggression than any other group (Figure 12), while girls without parental conflict had least aggression among them.

**Figure 12.**
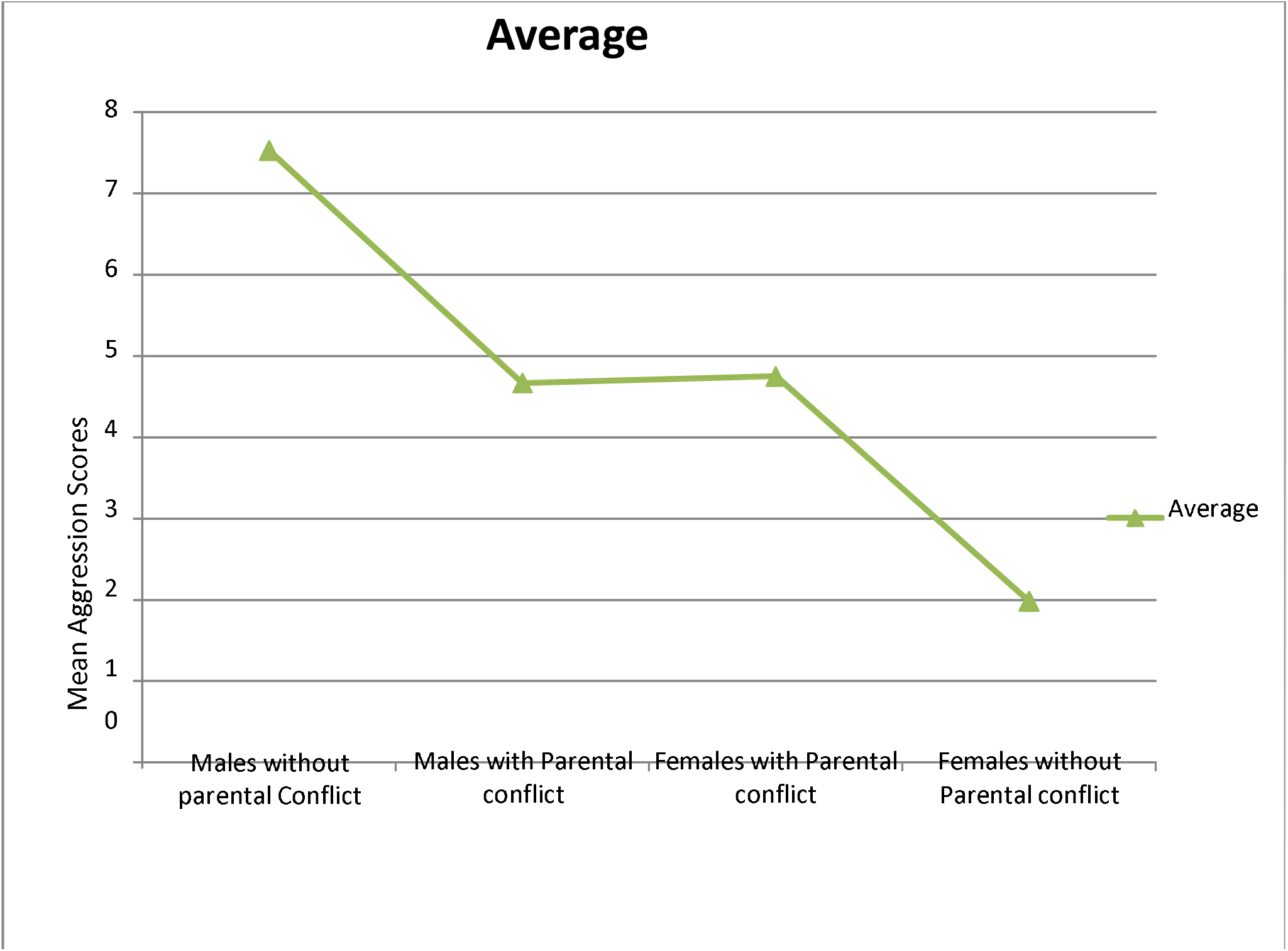
Comparison of males/females with/without parental conflict the results were analyzed by One-way ANOVA tests. It was found that aggression scores among males who didn’t perceive parental conflict were highest (mean=7.52) among all sub-groups, while it was minimum for females who didn’t perceive parental conflict (mean=1.98). (F=5.27, F_critical_ =2.69, p= 0.002). Since, F >F_critical_ at p=0.05 therefore, this difference is more than that expected by chance.

When data was analyzed for differences in aggression between those with physical abuse history, perceived parental conflict and combination of both the difference was not statistically significant (figure 13).

**Figure 13.**
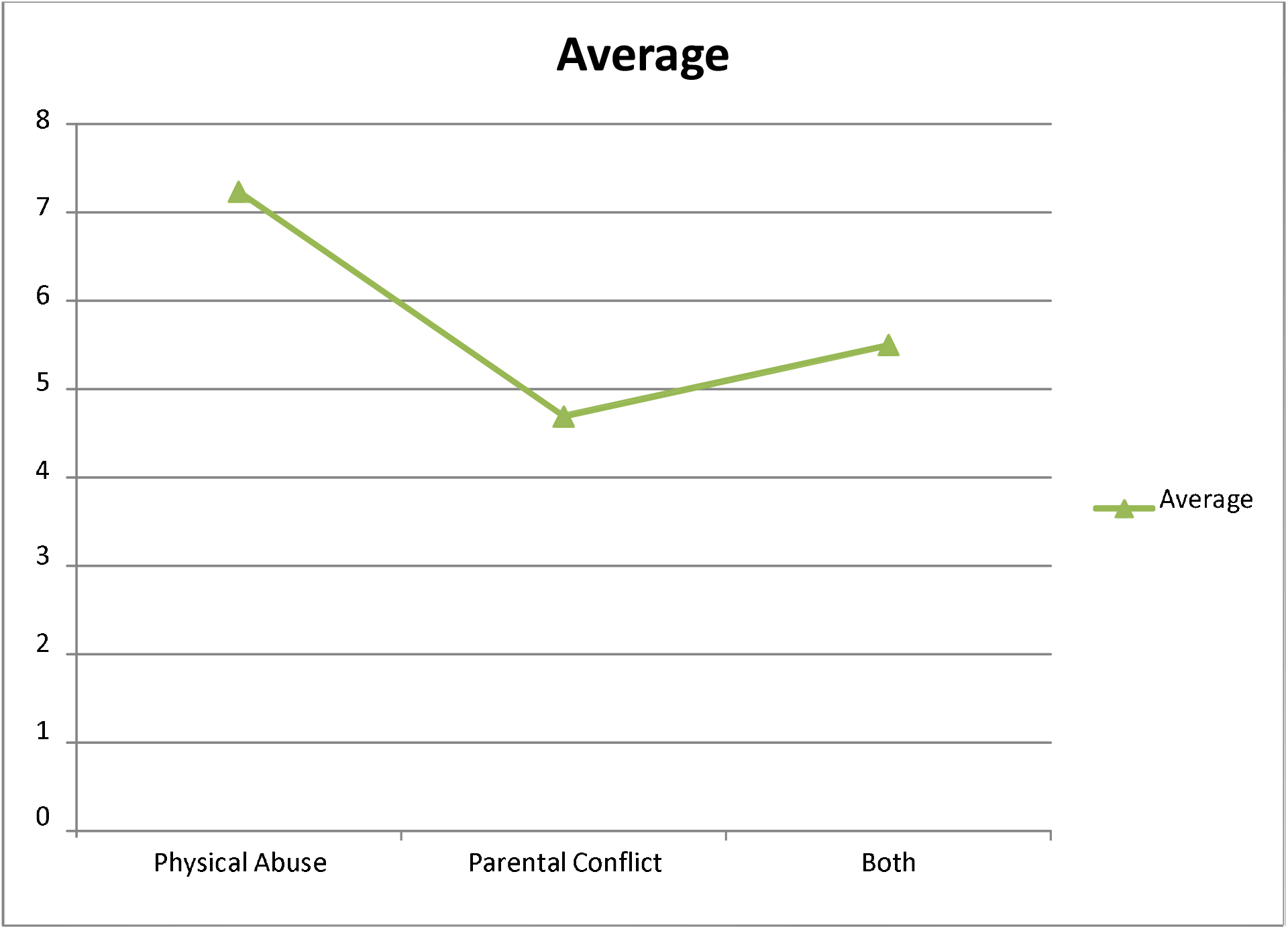
Comparison of aggression among those with physical abuse, parental conflict and combination of both using One-way ANOVA test. (F=0.545, F_critical_=3.18, p=.58). Since, F <F_critical_ at p = 0.05, therefore, difference between groups is **not** statistically significant.

Overall, analyses of data showed that among all the factors studied, aggression showed highest correlation with exposure to violence on TV for more than 2 hours/day (Figure 14).

**Figure 14.**
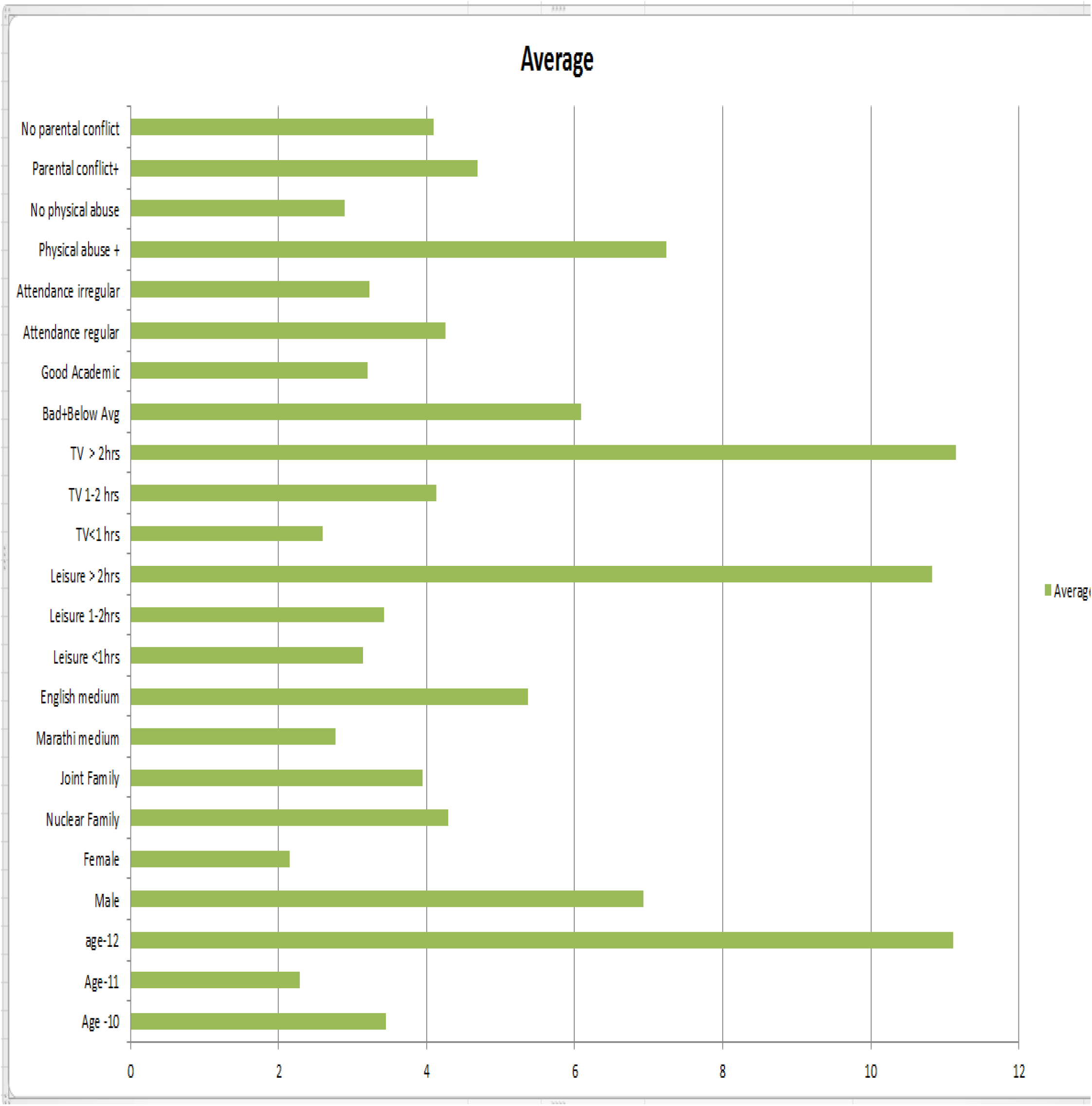
Y-axis represents the variables and sub-groups being studied and X-axis represents. On comparison of all the correlates it was found that those who were exposed to violence on TV for more than 2hrs/day had highest aggression scores of all (mean=11.15). Average aggression score of each variable (F=3.99, F_critical_=1.55, p=2.38 × 10^−9^).Since, F <F_critical_ at p = 0.05, therefore, difference between groups is not Statistically significant.

## DISCUSSION

Aggression in children and adolescents is a serious problem. It is associated with psychiatric disorders like conduct disorder, depression, school failure, substance abuse, child abuse and suicide. They are also at high risk for many other cardiovascular problems, cancer and brain damage.^20,21^ In our study, 4% of children from the age group of 10 - 12 years showed high levels of total aggression on the CAS-T scale. Moderate levels of aggression were shown by 8% of the children .Other studies have found the prevalence of aggression to vary from 4 to 14%. Frequent use of physical aggression by humans appears to reach its peak between 2 and 3 years of age.^22^ In the following years most children learn alternatives to physical aggression. Socialization of aggressive behavior during the preschool years should help prevent injuries throughout the life span.

It is important to control the aggression as these children victimize other children. These victims could also then be prone to psychological problems.^23^ Aggression also leads to teacher-child conflict which in turn can increase the aggression in children.^24^ Identification of the determinants is important to provide valuable information for teachers and health professionals so as to pay special attention to those at risk children.

Over the last decade researchers have found that boys are more aggressive thangirls.^25,26^ Similar findings were obtained in our study. The difference was statistically significant. Researchers have found that girls may be just as aggressive as boys when manipulative forms of behavior and spreading rumors, are included.^27^

Children studying in English medium schools had higher scores on CAS-T as compared to the vernacular medium. Cultural differences could exist between these 2 groups resulting in this significant increase in aggression among the English medium children. School and family environment also differ among these 2 groups. In India the changing family systems has resulted in an increase in the number of nuclear families. The joint or the extended family provided the child with more opportunities to express anger or hostility. Though no significant differences were found in the aggression seen in children coming from nuclear or joint families, the scores were higher in the nuclear family group.

Lower scores on CAS – T were found to be associated with a good academic school performance. Other studies have shown that students with low academic self-concept in achievement domains are more likely to be aggressive at school than those with high self-concept.^28^ It is also documented that aggression is related to a variety of poor academic outcomes such as: lowered academic performance, absenteeism and lower graduation rates. However, recent research has implicated only physical aggression as being more predictive of lower academic performance.^29^

The development of abnormally aggressive human behavior is complex and multifactorial. Aggressive patterns of behavior often begin early in life.^23^ Environmental factors influence the development of aggression by affecting children’s early relationships. In our study, higher scores positively correlated with the presence of physical abuse. Other studies have also found physical aggression in children with hostile or coercive parenting behavior.^22,26,30,31^ Early neglect and lower maternal warmth/sensitivity was also found to be associated with high aggression scores.^31,32^ The most commonly cited reasons for abuse included disobedience to parents, poor academic performance and quarrelling between parents.^33^

Thus parental conflict could lead to physical abuse causing aggression in children. Studies have also found that aggression is associated with parental conflict/depressive behavior or parental alcoholism.^32,34^ However in our study no significant association was seen. But when the groups were re-divided on basis of gender, there was more aggression present in boys who didn’t have parental conflict as compared to all other groups. While it was found that the girls who didn’t perceive parental conflict had least aggression. The child’s perception of the conflict may affect the child leading to more internalizing rather than externalizing behaviors. This could explain the lower mean scores obtained in children with presence of parental conflict. The study also showed that children who spent more than 2 hours per day in leisure time like play were found to higher scores on aggression. Other studies have also shown that children who spent excessive time in aggressive play or video games showed more aggression.^35^ Parents need to encourage children to spend their leisure time in non-aggressive activities.

On comparing the aggression scores based on the exposure to violence on TV significant higher scores were obtained in children who spent more than 2 hours in front of the TV watching such programs. Parents need to supervise what the child is exposed to. This would also include music and the internet. It is believed that repeated exposure to real-life and to entertainment violence may alter cognitive, affective, and behavioral processes, possibly leading to desensitization.^35^ There is consistent evidence that violent imagery in television, film and video, and computer games has substantial short-term effects on arousal, thoughts, and emotions, increasing the likelihood of aggressive or fearful behavior in younger children, especially in boys.^36^ By the time the average child leaves elementary school, that child will have viewed more than 8,000 murders and 100,000 other violent acts on network TV alone. It seems there is a bidirectional effect: More aggressive children prefer more violent TV, but TV violence increases their aggression even more.^37^ It was found in our study that when boys were exposed to violence on TV, they had highest mean of aggression(mean = 17.28) as compared to any other gender/TV exposure combination. Significantly, when all the variables of the study were assessed together using ANOVA test and later confirmed by Kruskal-Wallis Test (Nonparametric ANOVA) it was found that exposure of violence on TV for more than 2hrs/day had the highest aggression when compared to all other variables.

Thus the findings in the study indicate that preventive interventions should target families with high – risk profiles. Prevention programs should specifically target the parents’ control over their own aggression. Most prevention programs have targeted school age children but it is important to target these families in preschool years, as by 5-10 years aggression becomes a way of life. An early, cognitively based intervention may lead to reduced child aggression as a result of increased maternal social-emotional availability within the caregiving relationship.^38^ Violence and aggressive behavior are highly and disproportionately prevalent among school-aged urban minority youth, have a negative impact on academic achievement by adversely affecting cognition, school connectedness, and absenteeism, and effective practices are available for schools to address this problem.^39^ Also Preschool behavioral, emotional and motor problems, socioeconomic status, and family breakup are related to involvement in bullying at a later age.^40^

Interventions aimed at enhancing parent-child relationships or providing opportunities for alternative relationships with caring adult figures, may help to prevent abnormally aggressive behavioral outcome.^23^ Parental supervision of the child’s television viewing and play habits should form part of the interventions.

## CONCLUSIONS

Thus the study showed that high aggression levels were present in 4% of school going children of 10 -12 years.Moderate levels of aggression were present in 8% of the children. Thus about 12% of children require intervention for the aggressive tendencies.

Irrespective of gender, those children who watched violence on TV for more than 2 hours/day had highest aggression as compared to any other variable in the study. Among them theboys who watched violence on TV for more than 2 hours/day had more aggression than girls. Boys were found to be significantly more aggressive than girls in overall sample. Children from English medium schools were more aggressive than other medium. Aggression was also associated with lower academic performance and history of physical abuse. Perceived parental conflict was not significantly associated with increased aggression. However, males who didn’t perceive parental conflict had more aggression, which could be perhaps explained by child’s perception of the conflict, which may lead to more internalizing rather than externalizing behaviors.

Thus this study concludes that interventions to prevent aggression among children should include steps to supervise TV watching and limit the exposure of children to violence on TV. Awareness about the ill-effects of depicting violence in the media on this vulnerable population of children needs to create in society.Prevention programs should be targeted atthe high risk groups of children.Family environment modifications and changes in parenting styles at an early age could reduce aggressive behaviors in later years in these children.

## Data Availability

Data is avialable with the authors, and can be made avaiable on request

Since, a few columns in the above mentioned sample failed to pass the normality test (i.e. the sample was not distributed as per Gaussian curve) therefore, validity of ANOVA test was compromised.

Therefore, it was decided to perform **<u>Kruskal-Wallis Test (Nonparametric ANOVA)</u>**

The results of Kruskal-Wallis Test (Nonparametric ANOVA) were similar to ANOVA test

The **p value** obtained by Kruskal-Wallis Test (Nonparametric ANOVA) is **<u>0.001</u>** is considered very significant, and Variation among column medians is significantly greater than expected by chance.

